# Antiplatelet Therapy Following Spontaneous Coronary Artery Dissection: Systemic Review

**DOI:** 10.1101/2024.09.03.24312989

**Authors:** Huijun Edelyn Park, Leslie S. Cho, Natalia Fendrikova-Mahlay, Pulkit Chaudhury, Scott J. Cameron

## Abstract

**Background:** Spontaneous coronary artery dissection (SCAD) is an understudied cause of acute coronary syndrome (ACS), particularly in women. Heart muscle damage may result from spontaneous dissection of coronary arteries. There is no clear consensus regarding the optimum antiplatelet medication regimen and treatment duration for SCAD despite current American Heart Association (AHA) consensus guidelines recommending 12-month regimen of dual antiplatelet therapy (DAPT) consisting of a P2Y_12_ inhibitor and aspirin for patients following myocardial infarction (MI). The objective of this study was to evaluate the safety and effectiveness of DAPT compared to using a single antiplatelet therapy (SAPT) as part of the medical armamentarium to treat SCAD.

**Methods:** A comprehensive search of the literature was conducted to identify studies that examined SCAD outcomes including mortality, recurrence, and major adverse cardiovascular events (MACE) between 2000-2023 after antiplatelet therapy was administered. Based on the documentation in various studies, only 17 relevant studies were identified in which SAPT (primarily aspirin) and DAPT (aspirin combined with a P2Y_12_ inhibitor) were administered. Medications used in cardiovascular medicine that did not provide comprehensive data were excluded from the studies.

**Results:** DAPT treatment was associated with a poorer prognosis than SAPT 12 months after patients presented with SCAD. A key observation was the prevalence of antiplatelet treatment in SCAD patients, with DAPT prescribed for the majority of cases. At the 12-month time point, DAPT demonstrated had a higher rate of mortality (P = 0.0245), MACE (P = 0.0265), and angina admission rate (P = 0.071), as well as a higher rate of recurrent SCAD (P =0.0579) (Fig. 2). An increased incidence of non-fatal myocardial infarction or emergency percutaneous coronary intervention primarily drove these adverse outcomes.

**Conclusion:** In patients treated with antiplatelet therapy, adverse events that include unstable angina, mortality, and repeat revascularization were greater in patients with more aggressive antiplatelet therapy consisting of DAPT compared with these treated with SAPT.

## Introduction

Spontaneous coronary artery dissection (SCAD) is typically a manifestation in patients with non-inflammatory and non-atherosclerotic disease and can be a marker for an underlying collagen vascular disorder (1,2). Women are more prone to SCAD compared to men, potentially due to the differences in platelet activation between men and women both in healthy states and during myocardial infarction (6). A tear, also known as dissection, in the inner layer of coronary arteries can result in the creation of a false lumen in the wall of the artery, which may lead to a reduction or diversion of blood flow, and under-perfused myocardium that can precipitate myocardial infarction (MI) (2,4,5). While DAPT is considered the standard of care following MI in patients with atherosclerotic disease and acute plaque rupture, the appropriate antiplatelet therapy— if even required—following SCAD is not clear.

There is inherent risk to patients taking an antithrombotic agent following a diagnosis of SCAD. For example: platelet inhibition in the context of arterial dissection can lead to intramural hemorrhage—a potentially severe consequence of SCAD (4,6). While robust platelet inhibition reduces thrombotic risks associated with sub-intimal exposure and release of mediators that promote thrombosis, as well as shear-stress-mediated platelet activation, this also increases the risk of aggravating the already vulnerable vessel wall are increasing risk to the patient with unclear benefit (1, 2, 10).

Current Acute Coronary Syndrome (ACS) and American Heart Association (AHA) guidelines recommend administering a potent P2Y_12_ inhibitor for 12 months as part of a dual antiplatelet medication therapy (DAPT) (11, 12). Some studies found that clopidogrel, a P2Y_12_ receptor antagonist, reduces the risk of bleeding in SCAD compared with other antiplatelet drugs (3, 10, 11). There is a significant gap in the literature regarding antiplatelet therapy in SCAD, despite various treatment approaches being adopted in clinical practice (9–11). There are currently no evidence-based guidelines tailored to SCAD patients due to this lack of targeted research.

The major objective of this study was to compare DAPT with SAPT for medical management of SCAD following ACS. The passage discusses a study aimed at determining the best antiplatelet therapy for SCAD patients. Comparing an aggressive DAPT regimen with a more conservative SAPT approach to see which is more effective in managing SCAD, given the lack of clarity on the optimal treatment strategy. This research could help establish clearer treatment guidelines and improve patient outcomes.

## Methods

This systematic review evaluated the impact of antiplatelet therapy on SCAD patients in a comprehensive and structured manner. This review aimed to identify, assess, and synthesize relevant studies that compare the outcomes of DAPT and Single SAPT in patients with SCAD and followed PRSIMA reporting system algorithm.

### Search Strategy and Selection Criteria

A comprehensive literature search was conducted in the PubMed database, targeting a period between the years 2000 and 2023. The initial search criteria utilized specific keywords and phrases in the title and abstract, such as *(spontaneous coronary artery dissection [Title/Abstract]) AND (Antiplatelet therapy [Title/Abstract]), (spontaneous coronary artery dissection [Title/Abstract]) AND (DAPT [Title/Abstract]*”. This initial search resulted in the identification of 72 potential studies (**Table 1**.)

**Table 1:** Baseline Characteristics of the selected studies.

|  |  |  |  |  |  | Mortality (%) |  | MACE (%) |  | Recurrent SCAD (%) |  | Presence of Angina (%) |  |
| --- | --- | --- | --- | --- | --- | --- | --- | --- | --- | --- | --- | --- | --- |
|  | Study | Year | Design | N. of patients | Follow up | SAPT | DAPT | SAPT | DAPT | SAPT | DAPT | SAPT | DAPT |
| 1 | Mortensen *et al.*<sup>20</sup> | 2009 | Retrospective | 22 | 88 to 2932 days | 4 | 5 | 27 | 2 | 1 | 1 | NA | 22 |
| 2 | Alfonso *et al.*<sup>15</sup> | 2012 | Prospective | 45 | 2 years | 4 | 4 | 8 | 9 | 0 | 0 | 4 | 44 |
| 3 | Buja *et al.*<sup>16</sup> | 2013 | Retrospective | 38 | 17.4 months | 5 | 3 | 7 | 9 | NA | NA | NA | NA |
| 4 | Mc Grath-Cadel *et al.*<sup>30</sup> | 2016 | Retrospective | 40 | 16-month (from 6 months to over 4 years) | 0 | 0 | NA | 10 | 10 | 10 | NA | NA |
| 5 | Rogowski *et al.*<sup>14</sup> | 2017 | Retrospective | 64 | 4.5 years (1.8–8.4) years | NA | NA | NA | 5 | NA | NA | NA | NA |
| 6 | Chen *et al.*<sup>25</sup> | 2019 | Retrospective | 111 | 2.6 years | 0 | NA | 8 | 1 | 3 | 3 | 8 | 88 |
| 7 | Clare *et al.*<sup>17</sup> | 2019 | Retrospective | 208 | 4.7 years | 3 | 3 | 13 | 9 | 10 | 6 | NA | NA |
| 8 | Seidl *et al.*<sup>23</sup> | 2021 | Prospective | 105 | 4.5 years (1 to 3 months and 12 months) | 1 | 9 | 0 | 14 | 7 | 6 | 0 | NA |
| 9 | Cerrato *et al.*<sup>10</sup> | 2021 | Retrospective | 199 | 12months | 0 | 5 | 0 | 14 | NA | 6 | NA | 11 |
| 10 | Kim *et al.*<sup>29</sup> | 2021 | Retrospective | 13 | 121- 4125 days (11.3 years) | 0 | 0 | 7 | 7 | 7 | 7 | NA | NA |
| 11 | García-Guimaraes *et al.*<sup>28</sup> | 2021 | Retrospective | 318 | 6, 12 months, 5 years after the index event | NA | NA | 0 | 6 | NA | NA | NA | NA |
| 12 | Combaret *et al.*<sup>26</sup> | 2021 | Retrospective | 373 | one year | NA | NA | NA | 12.3 | NA | NA | NA | NA |
| 13 | Daoulah *et al.*<sup>18</sup> | 2021 | Retrospective | 83 | 18.8 months | NA | 12 | NA | NA | NA | NA | NA | NA |
| 14 | Saw *et al.*<sup>24</sup> | 2022 | Prospective | 750 | 1, 6, 12 months 3 years | 0 | 7 | NA | 14 | 3 | 3 | 1 | 1.06 |
| 15 | García-Guimaraes *et al.*<sup>19</sup> | 2021 | Retrospective | 389 | 29 months (follow-up ≥6 months) | NA | 2.5 | NA | 13 | NA | 2 | NA | 7 |
| 16 | de Sousa Almeida *et al.*<sup>27</sup> | 2023 | Retrospective | 36 | 40 months | NA | NA | 19 | 19 | NA | 14 | NA | 6 |
| 17 | Salamanca *et al.*<sup>21</sup> | 2023 | Prospective | 348 | 29 ± 11 months | 2 | 5 | 12 | 12 | 1 | 6 | NA | 6 |

### Inclusion Criteria

This search focused on English language articles, with an emphasis on meta-analyses, systematic reviews, and original research articles. Additionally, studies were screened based on the outcomes they reported, including mortality due to all causes, recurrent SCAD, angina, and major adverse cardiovascular events. The MACE process encompassed myocardial infarction, ischemic stroke, death, hospitalization, and bleeding complications. A total of 18 relevant studies were narrowed down through this process (**Fig 1**).

**Fig 1:**
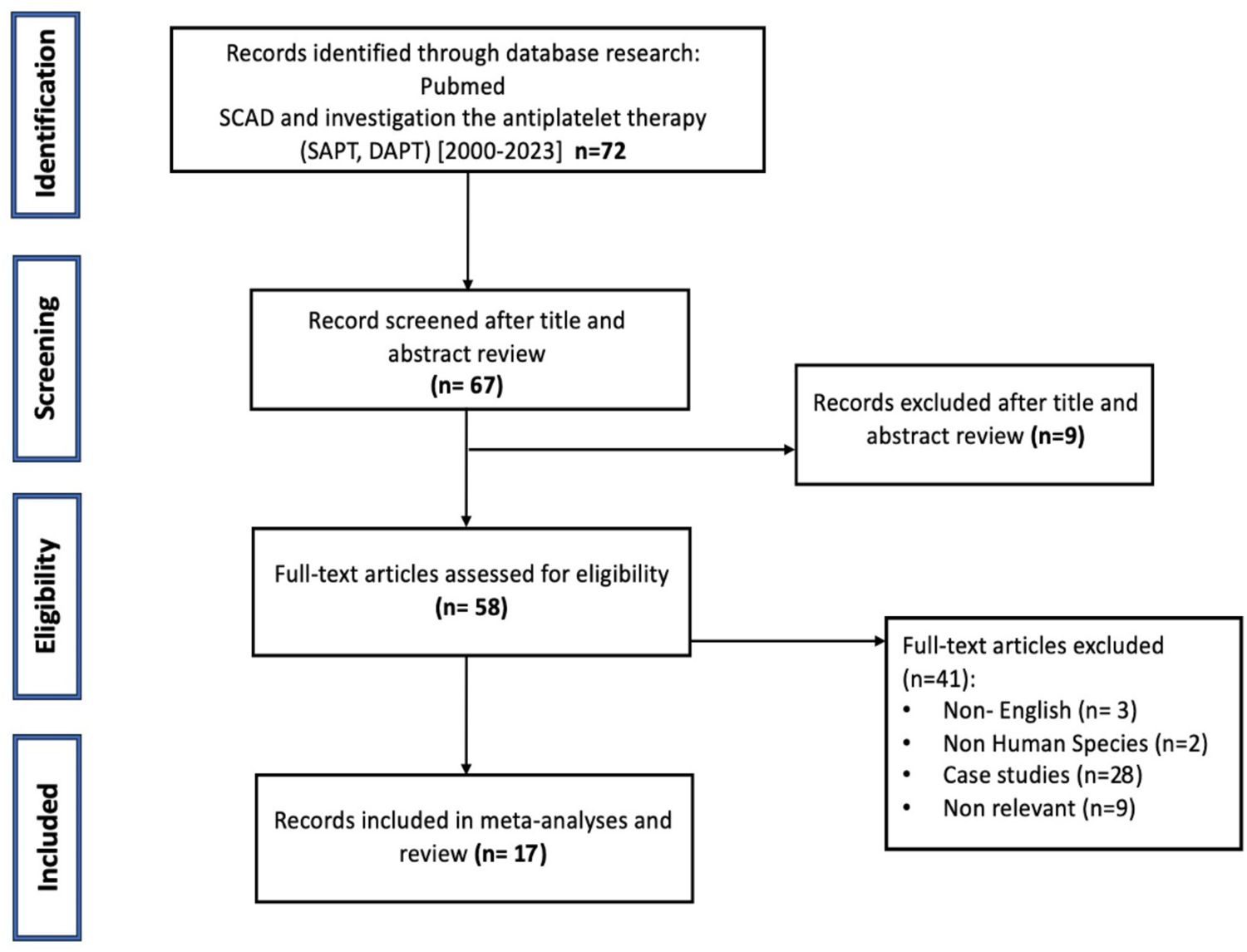
PRISMA flow diagram showing search strategy. Flow diagram showing the reasons for exclusion of articles from the study. The references included in the review are included as a separate list.

### Study Group Categorization

Study participants were categorized based on which antiplatelet regimen they were prescribed following diagnosis of SCAD. Two primary groups were identified: Single antiplatelet therapy (SAPT), initiated at the time of SCAD diagnosis, primarily comprising aspirin, and DAPT, which included aspirin in combination with a P2Y12 inhibitor (most commonly clopidogrel, ticagrelor, or prasugrel). Direct comparisons between the two therapeutic approaches were facilitated by this categorization. Additionally, we collected data on patient characteristics (such as age, gender, and ethnicity), type of SCAD, and treatment methods (including conservative management, percutaneous coronary intervention (PCI), or coronary artery bypass grafting (CABG)).

### Exclusion of Studies with Incomplete Data

Excluding studies without comprehensive data on cardiovascular medication use was a crucial part of the study selection process. In particular, studies that did not report the concurrent use of beta-blockers and statins, which are pivotal in the management of cardiovascular conditions, were excluded. As a result of this exclusion criterion, we ensured that the review only considered studies that offered a comprehensive view of patient management.

### Statistical Analysis

Continuous variables were assessed for normality using the Shapiro-Wilk test and found to be symmetric. Differences between groups was determined using the student’s t-test, or the Mann-Whitney U test for skewed data and a p value < 0.05 was considered significant.

## Results

Of the 72 studies identified to study SCAD, several were eliminated due to the absence of required data or elements required for study evaluation, leaving 17 studies available for evaluation. Heterogeneity analysis of reported outcomes revealed significant findings. A p-value less than 0.05 suggests that the observed effect or association is statistically significant. There was a statistically significant increase in mortality among DAPT-treated patients compared with SAPT-treated patients (P= 0.0245). There was also a significant increase in MACE in the DAPT group (P=0.0265 compared with SAPT). MACE included myocardial infarction, stroke, and the need for urgent cardiovascular interventions. In patients receiving DAPT following a diagnosis of SCAD, hospitalizations for angina were more frequent (P=0.0171). Lastly, there was a significant increase in the rate of recurrence in the SCAD group following DAPT (P=0.0579) compared to the SAPT group (Figure 2). Overall, DAPT was associated subsequently more adverse events in patients with SCAD, primarily due to myocardial infarction and unplanned percutaneous coronary interventions.

**Fig. 2.**
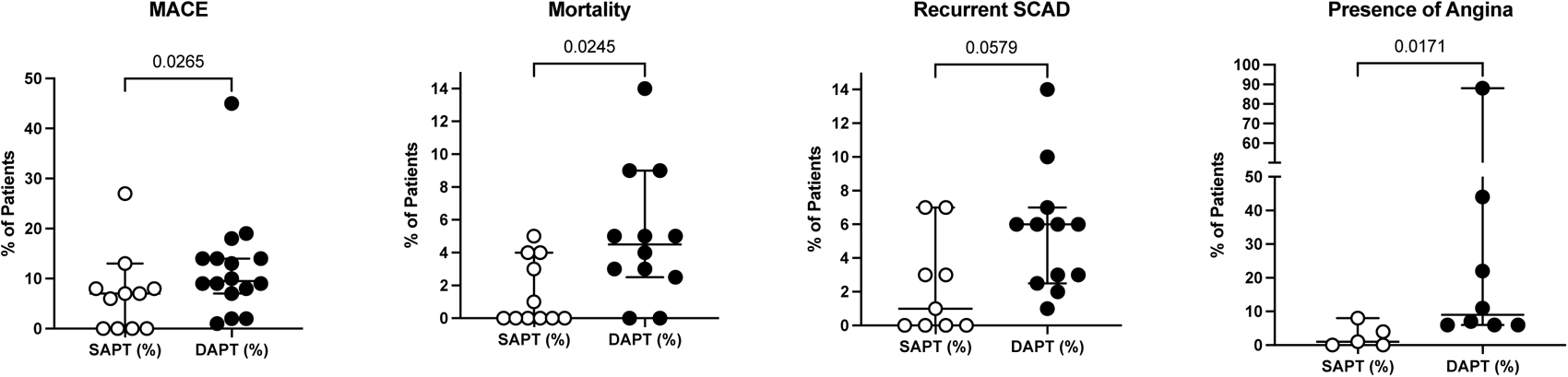
Outcomes in Patients with SCAD on Antiplatelet Therapy. Outcomes as a % of total patient population for each study were tabulated and data distribution was interrogated for normality and expressed as median ± 95% confidence interval. Group differences were assessed by the Mann-Whitney *U* test with the p-value as noted. The number of literature studies identified is indicated in parentheses. SAPT=Single Antiplatelet Therapy (white), DAPT= Dual Antiplatelet Therapy (black), MACE=Major Adverse Cardiovascular Events.

## Discussion

In this systematic review, we meticulously analyzed diverse studies to elucidate the clinical outcomes associated with SCAD, focusing on mortality, MACE, recurrence of SCAD (Re-SCAD), and present of angina. Specifically, mortality data were extracted from twelve studies (10, 15–21, 23–25, 30) providing invaluable insights into the fatal outcomes of SCAD. Furthermore, an in-depth evaluation of MACE was conducted through sixteen papers (10, 14–17, 19–21, 23–30), offering a comprehensive understanding of the major adverse cardiac events following SCAD. Recurrence of SCAD, a critical aspect of patient prognosis and management, was analyzed through twelve studies (10, 15, 17, 19–21, 23–25, 27, 29, 30). Additionally, eight studies (10, 15, 19–21, 24, 25, 27) were included to specifically address presence of angina, rounding out our examination of the vascular challenges posed by SCAD.

Patients with SCAD appear to benefit less from DAPT compared to SAPT, experiencing more frequent angina, recurrent SCAD, and increased mortality overall. In hospital unaccustomed to treating SCAD, DAPT may be administered for at least 12 months, a duration typically reserved for MI from atheroembolic disease or plaque rupture, despite the lack of high-quality outcomes data. Based on these findings, treatment protocols for SCAD need reevaluation taking into account the unique pathophysiological aspects of the disease and individual patient characteristics. In specific SCAD patient populations, a more conservative SAPT regimen may offer benefits, through further research is necessary to develop definitive guidelines.

Patients undergoing DAPT experienced higher rates of mortality, MACE, angina, and recurrent SCAD, as opposed to those on SAPT. The higher mortality rate observed in the DAPT group raises concerns regarding the safety and efficacy of this treatment approach for patients with SCAD. In addition, the recent meta-analysis revealed that in a meta regression analysis incorporating observational studies, the use of aspirin was associated with lower long-term rates of hospital admissions for angina, Additionally, a borderline association was observed between the use of DAPT and the recurrence rates of SCAD (35). Comparative analysis indicated that DAPT patients had a worse prognosis than SAPT patients. The outcome measures were statistically significant, indicating that the differences observed can probably be generalizable to SCAD patients. In the DAPT group, nonfatal myocardial infarctions and unplanned coronary interventions accounted for the majority of adverse outcomes.

In addition, the review findings challenge the current treatment protocols for SCAD, particularly the generalized inclination towards DAPT. A personalized treatment plan is needed for patients who experience SCAD, perhaps due to because of their variable response to antiplatelet therapy, but this should be formally investigated.

SCAD, often presents in young, otherwise healthy individuals, especially women, and requires a nuanced understanding of patient-specific factors, including hormonal influences, underlying vascular conditions, and unique characteristics of arterial dissection in SCAD in compared to atherosclerotic coronary artery disease (1,2, 11). To determine the appropriate antiplatelet regimen, it is important to recognize and respond to these differences.

Several factors might contribute to recurrent SCAD, in addition to comparing SAPT and DAPT. Beta-blockers are common medications prescribed for the treatment of cardiovascular conditions, including SCAD. Although in this review most patients treated with SAPT or DAPT are also prescribed beta-blockers, recurrent SCAD seems to be more prevalent among those on DAPT. Beta-blockers are known to reduce blood pressure, heart rate, and myocardial oxygen demand, which reduced angina. The finding that patients taking DAPT have a higher incidence of angina than those taking SAPT is interesting given that aspirin is known to decreased endothelial-derived dilating prostaglandins which precipitate arterial spasm (31), and arterial spasm may be underrecognized in patients following SCAD (32–34). However, there is some evidence suggesting that beta-blockers may also have potential adverse effects, in SCAD by causing hypotension and bradycardia (9). The exact mechanism of beta-blockers causing recurrent SCAD is unknown, but they may alter arterial tone and lead to vascular fragility, which may increase the risk of recurrence.

The association between beta-blocker use and recurrent SCAD is predominantly observed in patients receiving DAPT rather than SAPT. A potential interaction between beta-blockers and dual antiplatelet therapy warrants further investigation. A future study should investigate the specific mechanisms underlying this association and assess the overall risk-benefit profile of beta-blocker therapy in SCAD patients receiving different antiplatelet regimens.

As part of our study, we also tracked factors such as hypertension, diabetes, smoking status, dyslipidemia, stress-related SCAD, and pregnancy or hormone-related SCAD. However, there were no significant differences between patients treated with SAPT and those treated with DAPT in terms of these factors. Further, we monitored the use of angiotensin-converting enzyme (ACE) inhibitors, angiotensin II receptor blockers (ARBs), statins, percutaneous coronary intervention (PCI), and beta blockers for their potential influence on outcomes.

In spite of controlling for these variables, we consistently found that patients receiving DAPT had significantly higher rates of mortality, MACE, angina, and recurrent SCAD compared to those receiving SAPT. These findings indicate that variations in these factors cannot entirely account for the observed differences in outcomes between the two antiplatelet regimens. Further research is necessary to elucidate the mechanisms driving the adverse outcomes associated with DAPT in SCAD patients and to refine treatment strategies accordingly. Furthermore, only a limited number of studies have specifically examined recurrent SCAD cases. The limited number of studies examining recurrent SCAD emphasizes the complexity and rarity of the condition, as well as the difficulties associated with its diagnosis and treatment. Investing in research focusing on recurrent SCAD cases will contribute to a better understanding of the underlying mechanisms and risk factors associated with recurrence, as well as to the development of evidence-based treatment strategies for these patients.

To provide a deeper understanding of the optimal management of SCAD, more extensive studies, including randomized controlled trials of DAPT compared with SAPT, and SAPT consisting of aspirin alone of a P2Y_12_ receptor antagonist alone. A study of this type would be aimed at establishing definitive guidelines for the use of antiplatelet therapy in SCAD, potentially distinguishing it from the general protocol for patients following ACS.

## Limitation

Several limitations have been identified within the current body of research on SCAD, which is crucial for understanding the scope of our findings and guiding future research. Studies included in this review need to be revised regarding the size of their populations. This constraint potentially affects generalizability, highlighting the need for larger, more inclusive studies. To address this gap, we advocate the development of prospective, multicenter registries for patients with SCAD. SCAD registries would facilitate the detection of SCAD with greater accuracy and the collection of comprehensive patient data, improving the depth and breadth of information available for analysis. Across the studies reviewed, we observed considerable variation in the length and rationale of long-term follow-up. Most studies conducted follow-ups at 1, 6, and 12 months and annually after that for 3 to 10 years, but the average follow-up period was about five years. Despite concerted efforts to collect comprehensive follow-up data, it is possible that unrecognized cardiac or extracardiac vascular events or bleeding may occur. Thus, standardized follow-up protocols are essential to capture all relevant outcomes, thereby enhancing the reliability of long-term safety and efficacy data for patients with SCAD. We also examine the comparative outcomes of SART versus DAPT in patients with SCAD. As a result of the need for more high-quality data regarding the optimal duration and effectiveness of DAPT, it is necessary to reevaluate current treatment protocols. SCAD patients have a variety of characteristics and treatment responses, making it challenging to devise tailored treatment plans tailored to their individual needs. This underscores the importance of further research for developing tailor-made guidelines that consider the unique characteristics and treatment responses of patients with SCAD. Another limitation is the possibility of biases in selecting patients for Fibromuscular Dysplasia (FMD) screening. The Saw et al. (25) study indicated no difference in 3-year MACE between those screened for FMD and those not screened, although selection bias cannot be completely ruled out. Further emphasizing the need for comprehensive, unbiased research methodologies, this observation calls for a more nuanced understanding of FMD screening in the context of SCAD. These limitations are indicative of the complexity of SCAD, as well as the varied challenges faced by researchers in this area. A concerted effort is required to address these limitations by conducting large, methodologically sound studies with standardized protocols for diagnosis, follow-up, and treatment. SCAD can be better understood and improved for patients by overcoming these challenges.

## Conclusion

Exposing patients to DAPT following a diagnosis from MI as a consequence of SCAD appears to increase the risk of adverse cardiovascular events and mortality compared with SAPT. Mechanistic, and high-quality randomized clinical trials will be required to determine optimum antiplatelet for patients following a diagnosis of SCAD.

## Data Availability

All data produced in the present study are available upon reasonable request to the authors

## Acknowledgements

We would like to express our sincere gratitude to the following agencies for funding support, notably Pinkert family Foundation for their generous donation to Cleveland Clinic for SCAD research, and the National Institute of Health HL 158801-01 (to SJC).

---

**Supplemental data (Table 1).**

| Study | N. of patients | Mean follow up | Age (y) | Female (%) | Hypertension (%) | Diabetic (%) | Smokers (%) | Dislipidemia (%) | Stress related SCAD | ACE/ ATII (%) | Strains (%) | Beta-blockers (%) | PCI (%) | Bleeding events | Comments | Follow up |
| --- | --- | --- | --- | --- | --- | --- | --- | --- | --- | --- | --- | --- | --- | --- | --- | --- |
| Mortensen *et al.* <sup>20</sup> | 22 | 2,9 | 48 | 82 | 43 | 0 | 73 | 19 |  | 43 | 43 | 68 | 59 |  |  | 88 day to 2932 days |
| Alfonso *et al.* <sup>15</sup> | 45 | 2 | 53 | 58 | 33 | 11 | 62 | 38 |  | 53,3 |  | 80 | 26,7 |  |  | 2 years |
| Buja *et al.* <sup>16</sup> | 38 | 1,4 | 51 | 84 | 52,6 | 5,3 | 36,8 |  |  | 47,4 |  | 36 | 44,7 |  |  | 17.4 months |
| Mc Grath-Cadel *et al.* <sup>30</sup> | 40 | 3 | 45 | 95 | 18 | 5 | 8 | 10 | 22,5 | 40 | 27,5 | 52 | 30 |  |  | from 6 months to over 4 years |
| Rogowski *et al.* <sup>14</sup> | 64 |  | 53 | 94 |  |  |  |  |  |  |  | 55 |  | NA |  | 4.5 years (1.8–8.4) years |
| Chen *et al.* <sup>25</sup> | 111 | 3 | 48 | 92 | 28,8 | 7,2 | 12,6 | 37,8 | 12,6 | 63 | 81 | 88 | 48,6 |  |  | 2.6 years |
| Clare *et al.* <sup>17</sup> | 208 | 4,7 | 49 | 88 | 30,8 | 8,2 |  | 27,9 | 13,9 | 57,2 | 80,8 | 83 | 11,1 |  |  | 4.7 years |
| Seidl *et al.* <sup>23</sup> | 105 | 7,5 | 53 | 93 | 44 | 0 | 30 | 60 | 4,7 | 42 | 91 | 80 | 7 |  |  | 4.5 years (1 to 3 months and 12 months) |
| Cerrato *et al.* <sup>10</sup> | 199 | 1 | 52 | 88 | 39,7 | 3,5 | 26,1 | 37,2 | 49,5 |  | 71,3 | 78 | 0 | 2 (1) |  | 12months |
| Kim *et al.* <sup>29</sup> | 13 | 2,6 | 52 | 100 | 23,1 | 7,7 | 23,1 | 7,7 |  | 76,9 | 69,2 | 69 | 61,5 |  |  | 121- 4125 days (11.3 years) |
| García-Guimaraes *et al.* <sup>22</sup> | 318 |  | 53 | 88 |  |  |  |  |  |  |  | 79 |  | 2 (1) | In hospital follow up | 6 months and annually thereafter up to 5 |
|  |  |  |  |  |  |  |  |  |  |  |  |  |  |  |  | years after the index event |
| Combaret *et al.* <sup>23</sup> | 373 |  | 51 | 90 |  |  |  |  |  |  |  | NA |  | NA |  | one year |
| Daoulah *et al.* <sup>24</sup> | 83 |  | 44 | 51 |  |  |  |  |  |  |  | 89 |  | NA | Median f/up 18 months | 18.8 months |
| Saw *et al.* <sup>25</sup> | 750 | 3 | 51 | 88 | 32,1 | 6,9 | 11,6 | 20,3 | 60,1 |  |  | 84 | 14,1 |  |  | 1, 6, and 12 months and annually thereafter for 3 years |
| García-Guimaraes *et al.* <sup>19</sup> | 389 | 335 | 53 | 88 |  | 21 (5%) | 69 (18%) |  | 101 (26%) | 196 (51%) | 288 (75%) | 80 | 22 |  |  | 29 months (follow-up ≥6 months) |
| de Sousa Almeida *et al.* <sup>27</sup> | 36 | 3,4 | 51 | 94 | 53 | 6 | 31 | 39 | 8,3 | 61 | 72 | 78 |  |  |  | 40 months |
| Salamanca *et al.* <sup>21</sup> | 348 | 5 | 54 | 88 | 34 | 6 | 44 | 34 | 36,2 | 51 | 75 | 80 | 21 |  |  | 29 ± 11 months |

